# Mapping the Burden of COVID-19 in the United States

**DOI:** 10.1101/2020.04.05.20054700

**Authors:** Ian F. Miller, Alexander D. Becker, Bryan T. Grenfell, C. Jessica E. Metcalf

## Abstract

As of April ^5th^ 2020, SARS-CoV-2 has resulted in over 273,000 confirmed infections in the United States of America. Incidence continues to rise. As the epidemic threatens to overwhelm health care systems, identifying regions where the expected disease burden is likely to be high relative to the rest of the country is critical for enabling prudent and effective distribution of emergency resources. Across all global regions affected by the pandemic, an elevated risk of severe outcomes has consistently been observed in older age groups. Using age-specific mortality patterns in tandem with demographic data, we map a projection of the cumulative burden of COVID-19 and the associated cumulative burden on the healthcare system at the county-scale in the United States for a scenario in which 20% of the population of each county acquires infection. We identify regions that may be particularly impacted relative to the rest of the country, and observe a general trend that per capita disease burden and relative healthcare system demand may be highest away from major population centers.

## Introduction

The novel human coronavirus SARS-Cov-2 was first identified in December 2019 in Wuhan, China ^1^, and was observed in the United States shortly thereafter on January 7^th^ 2020 ^2^. The disease caused by SARS-Cov-2, termed COVID-2019, is both highly transmissible, with estimates of the basic reproductive number, R_0_, ranging from 1.4 - 6.49 ^3^, and clearly virulent, with an estimated overall case fatality rate of 1.4% ^4^, albeit this is highly dispersed across age groups. As of April 4^th^ 2020, the pathogen has spread globally, resulting in over 1,133,000 confirmed cases and killing over 62,000 individuals. Currently, the cumulative reported incidence of COVID-19 in the U.S. is the highest in the world, with over 273,000 confirmed cases ^5^.

As the COVID-19 epidemic expands within the U.S., a central focus of public health efforts will be limiting fatalities. A key driver of this outcome will be keeping the case burden of COVID-19 patients within the treatment capacity of the healthcare system. If the medical system is overwhelmed, case fatality rates are expected to increase, as severely ill COVID-19 patients will be unable to receive adequate care, or possibly any care at all, thereby exacerbating health outcomes. Critically ill patients may fare even worse; high mortality rates within this group would likely be further compounded by shortages of intensive care facilities and / or access to mechanical ventilation. Patients requiring care for non-COVID-19 reasons will also be indirectly affected by the health system’s inability to meet their needs.

The effective distribution of limited emergency medical resources, including personnel, protective equipment, and ventilators, to medical systems could help reduce the probability that specific hospitals will be overwhelmed by COVID-19 cases. Effectively distributing such resources requires information on the distribution of the burden of disease, and how that burden aligns with healthcare system capacity.

Several factors likely contribute to the heterogeneous distribution of COVID-19 burden across the U.S. The first of these is demography. A consistent trend in COVID-19 case distributions in the U.S. ^6^ and elsewhere ^7^ has been markedly higher incidence in older age groups (noting that observed cases do not necessarily reflect total infections). Several different mechanisms, including differential susceptibility or transmission potential between age classes, could potentially give rise to these patterns ^8^. However, differences in symptomatic infection rate between age groups has emerged as the most parsimonious explanation ^7,8^. Rates of hospitalization and intensive care unit admission have also been observed to be higher in older individuals ^9^. Spatial variation in age structure between counties, particularly the distribution of older individuals, may lead to differences in the per capita burden of disease between regions. Further, access to health care is another factor that could potentially affect the distribution of COVID-19 burden. Many rural areas of the U.S. may have insufficient ability to provide acute or critical care, or lack the capacity for both entirely. Residents of such areas may be at an increased risk for insufficient treatment. Finally, limited resources in rural areas may lead to an unexpected influx of cases to hospitals in population centers.

The temporal distribution of COVID-19 cases could also contribute to heterogeneity in disease burden. The magnitude and timing of the epidemic ‘peak’ is an obvious example -- the maximum demand for care determines the minimum healthcare system capacity necessary to provide adequate care. However, obtaining accurate predictions of the epidemic peak is a central challenge in emerging outbreaks due to limited, and often unreliable, data on incidence, and rapidly deployed and changing mitigation efforts. In the case of SARS-CoV-2, county-level variability in testing standards ^10,11^, Nonpharmaceutical Interventions (NPIs) ^12^, and timing of initial case introductions ^13^ limit our ability to produce accurate epidemic predictions at the weeks and months-ahead temporal resolution. While precise projections of peak disease burden would be highly illuminating for informing preparation efforts, such predictions are currently unreliable given the sparsity of data, particularly serological surveys. Uncertainty in region-specific transmission rates and the efficacy of mitigation efforts (e.g., timing, or absence, of shelter-in-place orders) further complicate long-term forecasting. In contrast, projections of cumulative disease burden are considerably more tractable. Such estimates will miss the nuance of the intensity and timing of outbreaks, yet their projection of the spatial footprint of disease burden contains core information relevant to resource distribution. In particular,\ comparing the total expected number of critical and severe infections against healthcare resources in each county in the U.S. allows for the identification of regions which may experience relatively high disease burden. Furthermore, analyzing simulations of multiple transmission scenarios (e.g., different contact patterns) makes it possible to identify regions of consistently high disease burden without needing to project an exact epidemic trajectory over time.

Here, we project the cumulative number of severe and critical COVID-19 cases in each county within the U.S. based on demographic data and age-specific risk factors for a scenario in which 20% of the population becomes infected. Using these projections, we calculate the healthcare system stress each county may experience as its own residents (and those from nearby counties with limited or non-existent medical resources) seek care. Finally, we map the expected burden of COVID-19 to identify the regions expected to experience the highest cumulative burden of disease in the long term.

## Results

We developed a modified age-stratified Susceptible-Exposed-Infected-Recovered (SEIR) epidemic model (based on the model of Davies et al. ^7^) to project the number of cases originating in each U.S. country. In this model, susceptible individuals (*S*) become infected in a density dependent fashion and enter the exposed (*E*) class, before eventually becoming either asymptomatically infected (*I*_*A*_) or ‘pre-clinically’ infected (e.g. symptomatic but not yet clinically presenting) (*I*_*P*_). The fraction of individuals who become pre-clinically infected rather than asymptomatically infected increases with age according to published estimates ^7^. Pre-symptomatic individuals eventually become symptomatic (*I*_*C*_). Asymptomatic and symptomatic individuals eventually recover with immunity to classes *R*_*A*_ and *R*_*S*_ respectively. All individuals in the infected classes (*I*_*A*_, *I*_*P*_, *I*_*C*_) are infectious; however, our model assumes that the relative infectiousness of asymptomatic individuals is reduced by a factor *b*_*a*_, and the relative infectiousness of clinical individuals is reduced by a factor *b*_*C*_ to account for the effects of case isolation and quarantine. This model is aimed specifically at projecting the age distribution of cases over a wide variety of transmission scenarios, and not at producing epidemiological forecasts. As such, we do not vary the components of our model linked to interventions, (e.g. contact rates) over time or by location

We chose to investigate a scenario in which 20% of the population in each county becomes infected. A 20% cumulative infection rate represents a pessimistic scenario for the short term (i.e., the next several months) but perhaps an optimistic scenario in the long term ^14^. We intentionally ignore spatial variation in the progression of the epidemic in order to simplify comparisons of disease burden between regions. As our aim is to provide general estimates about relative distribution of disease burden rather than to make precise predictions of case load over time, we sought to identify patterns of disease burden that are robust to different assumptions about the dynamics of epidemiological spread. To accomplish this, we varied our assumptions about the overall transmissibility of COVID-19, contact patterns between different age classes, and the contributions of clinically presenting individuals to transmission. For each set of assumptions, we simulated our model for each county in the U.S. using demography data from the 2018 American Community Survey, and extracted the number of individuals in each age class who had become symptomatically infected by the time the cumulative population infection rate had reached 20%. In the main text, we present detailed results for the most optimistic scenario and most pessimistic scenario. Summarized results for 25 other scenarios are presented in the *Supplementary Materials*. In the ‘optimistic’ scenario (transmission, *R*_*0*_ = 2, mixing structure, *Θ* = 1, reduction in relative infectivity of clinically presenting individuals, *b*_*C*,_ = 0.1, see *Methods* for further details) transmission is relatively slow, contact patterns exhibit a strong age structure, and clinically presenting individuals are effectively quarantined. In contrast, the ‘pessimistic scenario’ (*R*_*0*_ = 6, *Θ* = 0, *b*_*C*_ = 1) is characterized by high transmission, well-mixed contact patterns, and ineffective quarantine.

Using our projections of cumulative symptomatic infections, we further estimated the number of severe cases (i.e., requiring hospitalization) and critical cases (i.e., requiring intensive care) using published rates of these outcomes for various age classes ^9^. We found that in all transmission scenarios, the areas with high relative burdens of hospitalizations and intensive care unit (ICU) admissions were those with high population density (See Figure 1 A, Figure 2 A, D, and Figure 3 A, D). However, we observed the opposite pattern for the per capita burden of hospitalizations and ICU admissions, which were distributed heterogeneously and were generally highest away from major population centers. Due to the positive correlation between age and disease severity, the areas with the highest per capita burden were those with older populations (See Figure 1 B, Figure 2 B, E, and Figure 3 B, E,). Correspondingly, we identified a positive correlation between both hospitalization and ICU admission rates with age across all counties. We further found that older age classes were even more disproportionately affected in the ‘pessimistic’ transmission scenario that assumed no age assortative contact patterns (See Figure 2 G, Figure 3 G). Despite this, we found that the sets of counties with very high projected burdens of per capita hospitalizations and ICU admissions remained similar across different transmission scenarios. Indeed, of the 315 counties at or above the 90% quantile of per capita hospitalization in the ‘optimistic’ transmission scenario (see Figure 1 legend), 244 were also at or above this quantile in the ‘pessimistic’ scenario. Of the 315 counties at or above the 90% quantile of per capita ICU admissions in the ‘optimistic’ transmission scenario, 258 were also at or above this quantile in the ‘pessimistic’ scenario.

**Figure 1:**
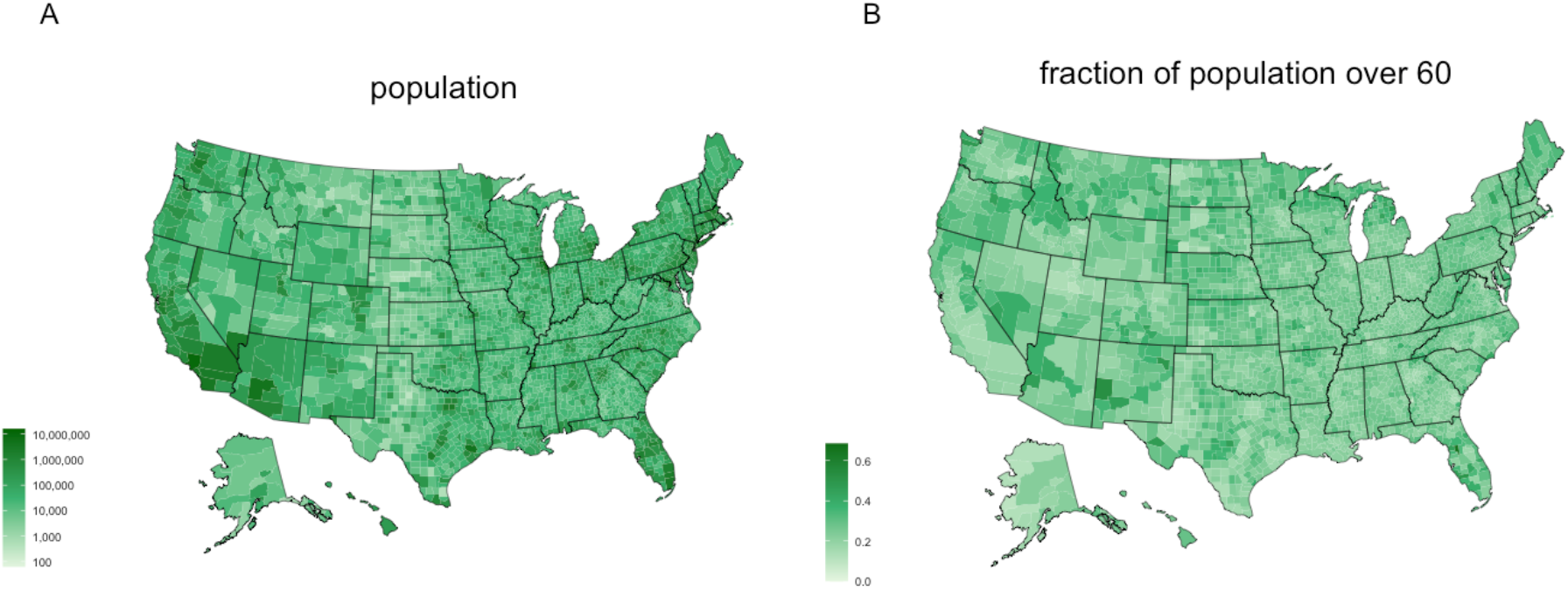
Population characteristics of the U.S. Panel A shows the population of each county. Panel B shows the fraction of individuals within each county over the age of 60.

**Figure 2:**
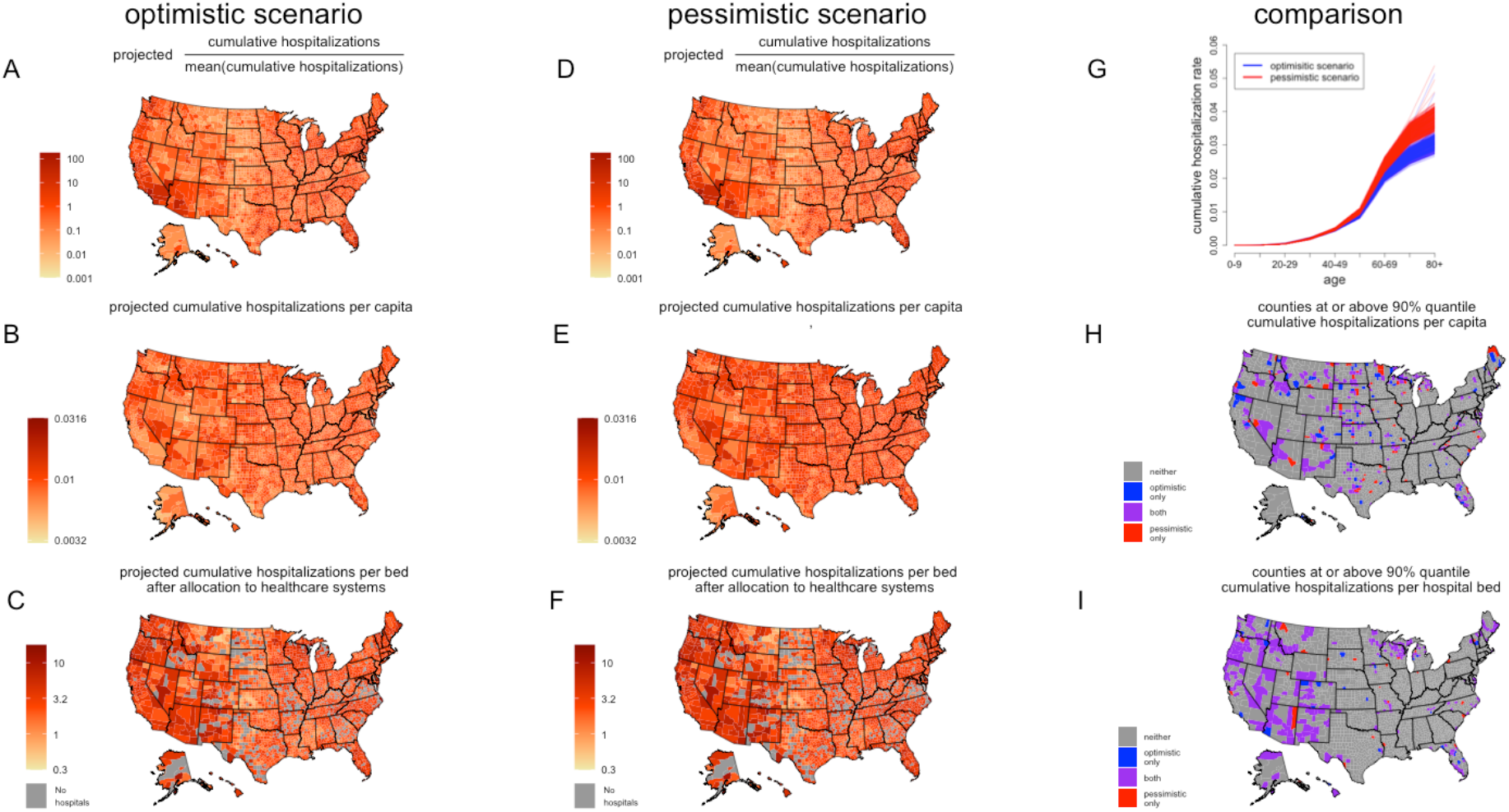
Burden of hospitalizations in the US. We mapped the burden of COVID-19 hospitalizations (assuming a 20% cumulative infection rate) in the U.S. Panels A, B, and C show results for the ‘optimistic’ scenario, and panels D, E, and F show results for the ‘pessimistic’ scenario. A and D show the relative number of hospitalizations in each county. B and E show the number of projected hospitalizations per capita in each county. In A, B, D, and E cases have not yet been allocated to healthcare systems. C and F show the cumulative number of hospitalizations per hospital bed after cases have been allocated to healthcare systems. G shows the percentage of each age class infected in each transmission scenario. Each of the 315 lines for each transmission scenario represents a different county. H and I show the counties estimated to be in the 90% quantile of hospitalizations per capita and hospitalizations per hospital bed (after case allocation). Colors in H and I indicate whether the counties were estimated to be in the 90% quantile in the optimistic scenario, the pessimistic scenario, both, or neither. A high-resolution version of this figure is provided in the supplementary materials.

**Figure 3:**
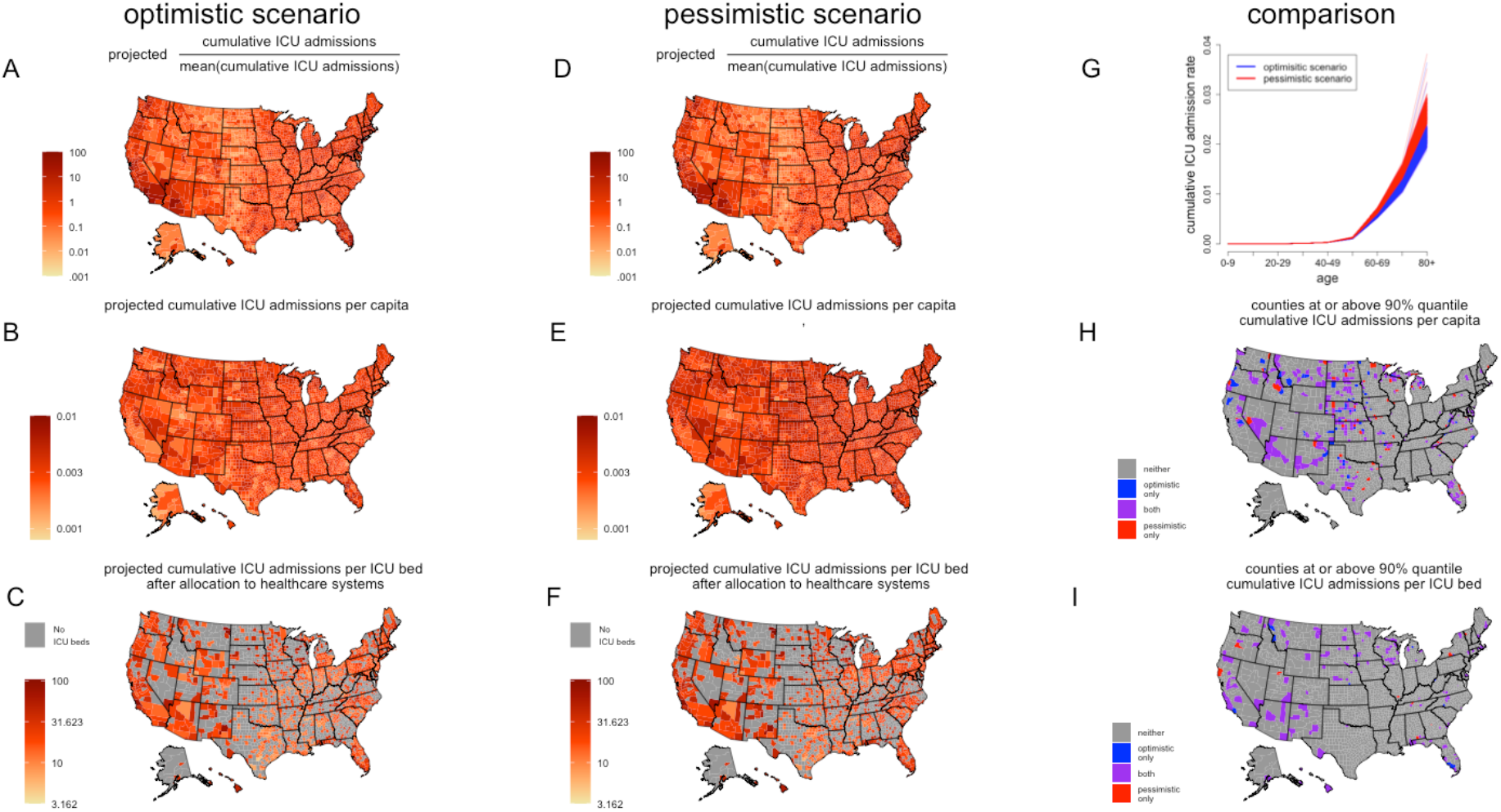
Burden of ICU admissions in the US. Figure 3 shows the projected burden of COVID-19 in terms of ICU admissions. Scenarios and subplots are the same as in figure 2. A high-resolution version of this plot is provided in the supplementary materials.

Next, we sought to identify how patterns of case burden aligned with healthcare system capacity. We distributed cases to healthcare systems within and outside of their county of origin based on an allocation algorithm (see *Methods*). In brief, this algorithm distributes severe and critical cases based on relative distance and the relative capacity of healthcare systems to provide care (quantified as the number of hospital beds and ICU beds respectively). The vast majority of cases originating from within a county with substantial medical resources stay within that county. Most severe and critical cases originating from within a county with few hospitals or ICU beds are allocated to nearby counties with greater care capacity. All severe or critical cases originating in a county that lacks the capacity to provide appropriate care entirely are distributed to nearby counties.

The maps of relative hospitalizations per bed (See Figure 2 C, F) and relative ICU admissions per bed (See Figure 3 C, F) indicate which counties are expected to experience a relatively higher burden of disease relative to medical resources. Several regions have higher concentrations of these counties, including much of the western United states, the northern Midwest, Florida, and northern New England. These patterns appear to be robust to assumptions about transmission rates and age-specific mixing patterns. The ‘optimistic’ and ‘pessimistic’ transmission scenarios each identified 248 countries as being at or above the 90% quantile of cumulative hospitalizations per hospital bed, and 223 counties were identified in both transmission scenarios. In the case of ICU admissions per bed, 125 of the 136 counties identified as being at or above the 90% quantile were the same for both transmission scenarios.

## Discussion

Even with unprecedented efforts to speed the development of a vaccine ^15^, it is unlikely that a pharmaceutical intervention against COVID-19 will become available in the near future. SARS-Cov-2 transmission is expected to continue over the coming months and affect virtually every locality in the U.S. The central aim of our analyses is to identify counties that consistently emerge as being likely to experience a relatively large burden of disease (across a range of assumptions about transmission) on their population and healthcare systems to inform and urge both preparedness and the allocation of emergency medical resources. We identified several regions in need of support, including much of the western portion of the country, the northern Midwest, Florida, and northern New England. At a finer geographical scale, our results suggest considerable rural-urban inequity, with the per capita burden of disease being highest away from major population centers.

Before even considering the increased case burden these more rural places are projected to experience relative to the rest of the U.S., it is evident that hospitals, and to a greater extent, hospitals with the capacity to provide intensive care, are unevenly distributed. Many regions have limited, or even no, facilities with the ability to provide the type of acute or critical care needed to treat COVID-19. Case fatality rates in these regions may rise above the national average if people are unable to access care. Bolstering the capacity of rural health systems and ensuring equitable access to care should be central goals of COVID-19 management strategies. It is important to note that while the healthcare systems of major population centers aren’t identified as weak spots in our analysis, they do service a much larger number of people. Given the magnitude of the consequences of their potential failure, they should, of course, remain a priority for preparedness efforts.

Our findings are robust to different assumptions about transmission patterns. However, it is imperative that they be interpreted in the context of our methods. We were deliberately conservative in not considering the impact of potential therapeutics and vaccines. Our results only underscore the urgency of developing these interventions. We specifically did not attempt to predict the epidemic peak timing or magnitude. Given the time-invariant scenario we model (i.e., 20% of the population acquires infection), it is likely that our projections will not match future observed patterns of disease burden in the short term as many regions are still in the early phases of their epidemics. However, our results provide a first best guess as to the expected patterns of burden in the long term rooted in basic features of demography and health system capacity. Notably, we did not consider how other factors, such as comorbidities ^16^ (e.g. hypertension, pulmonary disease) or socioeconomic status ^17^, might result in increased disease burden in certain regions. Incorporating such factors into mathematical models and their forecasts is an essential area of future research. Finally, we urge public health officials using our results to carefully consider location-specific details and nuances not explicitly included in our analyses when planning their response.

In conclusion, we have identified areas in the U.S. expected to be particularly heavily affected by COVID-19. Our findings suggest assisting hospitals and communities away from major population centers will be crucial as we aim to mitigate the consequences of the ongoing COVID-19 epidemic.

## Data Availability

Code and data associated with the analyses are available via github.

https://github.com/ianfmiller/covid19-burden-mapping

## Acknowledgements

I.F.M and A.D.B. acknowledge funding from National Science Foundation Graduate Research Fellowships.

## Author contributions

I.F.M., A.D.B., and C.J.E.M. conceived of the study. I.F.M and A.D.B. conducted the analyses. All authors contributed to the writing and revision of the manuscript.

## Methods

### Data

We obtained counts of the number of individuals in 10 year age bins in each U.S. county from the 2018 American Community Survey available from the United States Census Bureau. We define the set of age categories as *G={0-9, 10-19*,…*70-79, 80+}*. We obtained data on hospital location and bed number from the American Hospital Association 2018 annual survey. We used the calculated total of all beds for each hospital to represent the number of hospital beds, and the number of adult medical/surgical intensive care beds to represent the number of intensive care unit beds. We aggregated hospital and ICU bed data by county in accordance with American Hospital Association data use policy.

### Mechanistic models

We developed an age stratified mechanistic epidemiological model based on that of Davies et al. ^7^ that follows a susceptible-exposed-infected-recovered (SEIR) framework. This model assumes no births or deaths. The subscript *i* denotes the index of the age strata. The parameter *r*_*i*_ denotes the rate of symptomatic infection for age class *G*_*i*_. We set values for *r*_*i*_ according to the approximation for Wuhan, China from Davies et al.

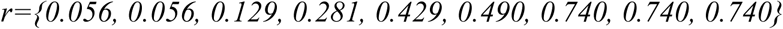

The infected class is decomposed into asymptomatic (*I*_*A*_), symptomatic, pre-clinical (*I*_*P*_), and symptomatic, clinical (*I*_*C*_) classes order to reflect relevant aspects of SARS-Cov-2 epidemiology, namely that not all infected individuals show symptoms, and individuals are frequently quarantined upon presenting symptoms. We also decomposed the recovered class into separate compartments for those recovered from symptomatic infection, *R*_*S*_, and those recovered from asymptomatic infection, *R*_*A*_, in order to simplify calculations of total symptomatic and asymptomatic cases. This model framework allows us to impose assumptions about the infectivity of asymptomatic individuals and clinically symptomatic individuals (*b*_*A*_ and *b*_*C*_ respectively) relative to the infected class likely responsible for the bulk of transmission (*I*_*P*_).

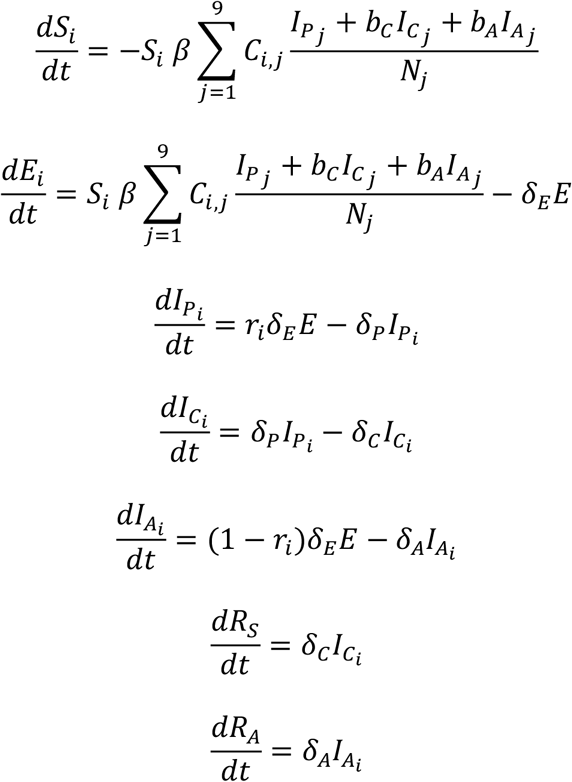

Here, C is the contact matrix, whose entries *C*_*i,j*_ correspond to the mean number of contacts between individuals in the *i*-th and *j*-th age classes of *G, δ* parameters determine the mean amount of time that individuals spend in each class, and *β* is the transmission parameter.

We used this model to simulate a wide range of plausible epidemiological scenarios. Specifically, we considered values for *b*_*C*_ in {0.1, 0.5, 1}, values for *R*_*0*_ in {2, 4, 6}, and values for the degree of homogeneous mixing, in {0, 0.5, 1}. In the sections below, we describe how we use to construct the contact matrix *C*. We set the values of the following model parameters according to published estimates^7,18,19^ 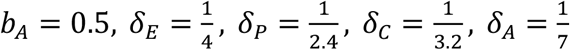 After constructing *C* and fixing these variables, we used numerical methods combined with the next-generation matrix approach^20^ to calculate the value for that corresponds to the value R_0_ we wished to assume for each scenario.

### Rescaling contact matrix

We used the ‘*socialmixr’* ^21^ R package to retrieve the UK contact matrix from the POLYMOD study^22^ with contacts binned according to the following age categories: {0-9,10-19,…60-69,70+}. We term this matrix *A*. No finer resolution was available for contacts involving individuals over the age of 70. However, in order to account for differences between individuals in the 70-79 and 80+ age groups in terms of relevant COVID-19 parameters, we synthesized a new matrix, *B*, that includes contacts for individuals in the 70-79 and 80+ age classes:

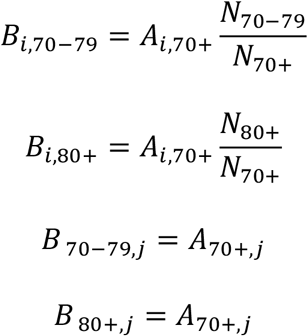

Here *N*_*x*_ is the number of individuals in the entire U.S. in age class x.

Next, we constructed the contact matrix used in our model *C* by rescaling *B* to reflect our assumptions about mixing patterns.

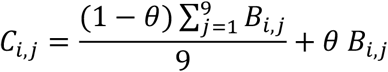

The quantity *θ* represents the degree of homogeneous mixing. When *θ* = 1, contact patterns are identical to the POLYMOD contact patterns. When *θ* = 0, contact rates are homogenous across age classes. Values of *θ* between 0 and 1 correspond to mixing patterns intermediate between the POLYMOD and homogenous scenarios. This rescaling procedure preserves the total number of contacts that each age class experiences while changing the identity of those contacts.

### Model simulation

For each scenario in each county, we used the following conditions to initiate the model:

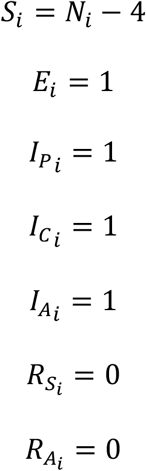

The number of individuals within each age class for the county of interest is *N*_*i*_.

We then simulated the model in R, using the ‘ode’ function in the ‘*deSolve’* package^23^ with the ‘*lsoda’* integrator and a step size of 0.25. We truncated the simulation when

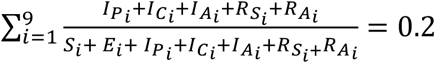

and then extracted the number of individuals in each age stratified compartment.

### Case Estimation

We calculated the total number of symptomatic infections in each age class by the time the cumulative infection rate reaches 20% as 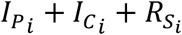 at the end of the simulation. We then calculated the number of hospitalizations in each age class by multiplying the number of symptomatic infections in each age class by age stratified estimates^9^ of hospitalization rates for symptomatic cases: {0.001, 0.003, 0.012, 0.032, 0.049, 0.102, 0.166, 0.243, 0.273}

We then calculated the number of ICU admissions in each age class by multiplying the number of hospitalizations by age stratified estimates^9^ of the rate of ICU admission given hospitalization: {0.05, 0.05, 0.05, 0.063, 0.122, 0.274, 0.432, 0.709}.

### Case Distribution

We distributed cases originating in a given county to the healthcare systems of that county and other counties using the following algorithm.

- Let the county of origin be denoted as *c*_I_ and the potential destination counties as *c*_I_,…, *c*_\_
- Let the distances between the center of population of the county of *c*_I_ and each potential destination county *c*_i_ be *d*_I,i_

○ We obtained the latitude and longitude of the center of population for each county from publicly available data from the 2010 U.S. census, and calculated pairwise distances between counties using R package ‘*geosphere’* ^24^.
- We next removed all destination counties with *d*_I,i_ > 400km.
- We calculated a distance weight, y_i_, for each remaining potential destination county as

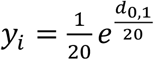
- We calculated a bed weight, *z*_i_, for each county as the number of total hospital beds in *c*_i_.
- For projections involving ICU admissions, we used the number of ICU beds instead of the number of hospital beds.
- We then calculated a composite weight, *w*_i_, for each county as 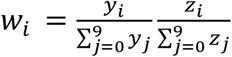
- Lastly, cases originating in *c*_I_ were then distributed to counties *c*_0_,…, *c*_N_
- proportion to 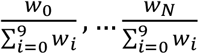

